# Landscape of Gene Mutation in Synovium of Patients with Rheumatoid Arthritis

**DOI:** 10.1101/2020.04.14.20064592

**Authors:** Jun Inamo

**Author notes:** Correspondence to: Jun Inamo, M.D., Address: 35 Shinanomachi, Shinjuku-ku, Tokyo160-8582, Japan.

## Abstract

**Objective:** To investigate accumulating single nucleotide variants (SNVs) in synovium of patients with early, pre-treatment rheumatoid arthritis (RA).

**Methods:** I leveraged RNA-seq dataset derived from patients with RA deposited in database. To capture synovium-specific mutations, I applied standardized SNVs-calling method to paired synovium and blood samples per individual.

**Results:** De novo mutational signatures constructed by non-negative matrix factorization reflected continuous relationship of synovial histology, namely lymphoid-myeloid (Lymphoid), diffuse myeloid (Myeloid) and pauci-immune fibroid (Fibroid). Using COSMIC signature, defective DNA damage repair-associated mutational signature was associated with the level of inflammation in synovitis. Further, pathway analysis based on specific genes harboring SNVs in each histological group represent different types of synovitis, with differing pathogenic process and inflammatory environment.

**Conclusion:** This data will promote our understandings about pathogenic status of synovitis, and suggests evidence that stratified treatments would be optimal according to histological subgroup.

## Introduction

Single nucleotide variants (SNVs) could be certainly propagated from progenitors to daughter cells during DNA replication and thus have been commonly used to reveal the characteristics of cancer cells [1]. Not only tumorigenesis, there is also increasing evidence that the genetic architecture affect susceptibility to rheumatoid arthritis (RA) [2]. In addition to germline mutation, somatic cells certainly acquire genetic mutations with ageing and environmental factors [3]. Recent study reported a unique pattern of somatic mutagenesis in the inflamed tissue and it may be causally linked to the inflammatory process [4]. SNVs could be detected from the whole-genome sequencing data (WGS) or whole exome sequencing (WES), and then be utilized to infer impact on nature of cell populations. While the next-generation sequencing (NGS) of RNA samples (RNA-seq) is designed to quantify gene expression, it also provides opportunities to investigate gene or transcript differences in lesion at a nucleotide level. Indeed, previous study demonstrated usefulness of RNA-seq to detect genomic variants [5]. However, regarding to autoimmune diseases, association between SNVs obtained by RNA-seq at lesion and phenotype hasn’t been largely known.

Here, SNVs accumulating in synovium were identified in patients with early, pre-treatment RA. Further, signature of SNVs was different according to subgroups of synovial histology, namely lymphoid-myeloid (Lymphoid), diffuse myeloid (Myeloid) and pauci-immune fibroid (Fibroid).

## Materials and Methods

### Subjects

RNA-seq dataset was downloaded from ArrayExpress under Accession code E-MTAB-6141. The details of subjects were available in previous report [6]. Briefly, I leveraged RNA-seq dataset derived from patients with early, pre-treatment RA (Supplementary Table 1; Lymphoid, n=11; Myeloid, n=5; Fibroid, n=3). Histological subgroup was based on histology scores; synovial samples were classified as Lymphoid (CD20 B cell aggregate rich), Myeloid (CD68 rich in the lining or sublining layer but poor in B cells), or Fibroid (paucity of immune-inflammatory cell infiltrate). RNA from blood and synovium samples were extracted and sequenced by standardized methods as described in previous report [6].

### Single nucleotide variants-calling methods

The multi-step low-quality read-pair filtering was as described in the GATK Best Practices workflow for SNV calling on RNAseq data (Figure 1) [7]. First, downloaded fastq files were assessed for quality using the reports generated by FastQC (v.0.11.9). Subsequently, low-quality regions and adapters were trimmed using Trim Galore (v.0.6.5) (threshold of phred scores was 30). Filtered reads were then mapped onto the human reference genome version hg38 using STAR’s two-pass mode (STAR, 2.7.3a). Sorting, read groups assignment and duplicates marking were performed by Picard toolkit (v.2.21.9). Generated reads were passed to the Genome Analysis Toolkit (GATK, v.4.1.4.1). After reads were cleaned by MergeBamAlignment and MarkDuplicates tools, reformat of the alignments was conducted by SplitNCigarReads tool, which splits reads with N in the cigar into multiple supplementary alignments and hard clips mismatching overhangs. After base quality recalibration, SNVs were called using the HaplotypeCaller tool. Variants were annotated using SnpEff (v.4.3). Raw variants were filtered by several threshholds (Figure 1). For each dataset, a hard filtering step was applied (options—clusterSize 3—clusterWindowSize 35—filterExpression “FS > 60.0”—filterName FS—filterExpression “QD < 2.0”) to filter out SNVs in clusters (more than 3 SNVs in a window of 35 bases), and variants with QualByDepth (QD) < 2.0 and FisherStrand (FS) > 60 accounting for variant quality and strand bias. After subtracting SNVs from list of SNVs of each sample, SNVs which were also detected in the paired blood sample per individual were removed to focus on specific SNVs in synovium. To extract distinctively conservative SNVs in each subgroup of synovial histology (Lymphoid, Myeloid and Fibroid), only common SNVs in each subgroup were proceeded to further analysis. List of SNVs identified by above pipeline are available in Supplementary Table 2-5 (https://figshare.com/articles/Supplementary_Tables/11876553).

**Figure 1.**
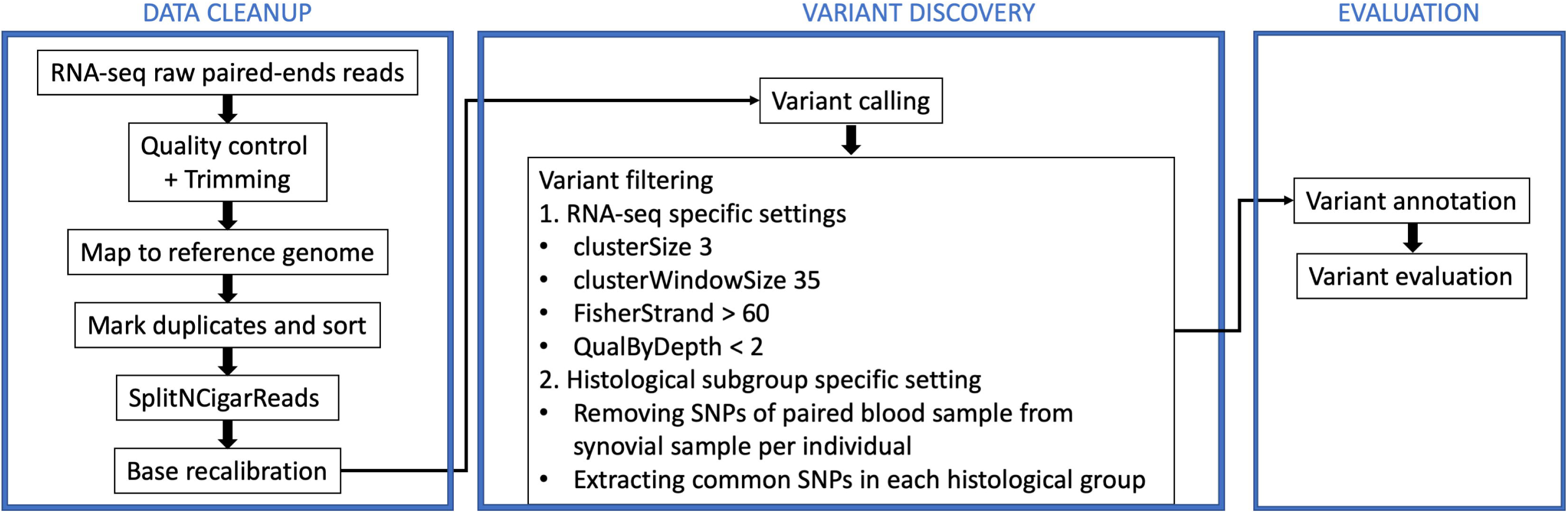
Work flow in this study. Three-step approach was used according to The Genome Analysis Toolkit recommendation for calling variants from RNA-seq.

### Mutational signatures

Filtered variants were used to visualize a multitude of mutational patterns in base substitution using the R package MutationalPatterns (v.3.10). The profile of each mutational signature is displayed using the six substitution subtypes: C>A, C>G, C>T, T>A, T>C, and T>G. Further, each of the substitutions is examined by incorporating information on the bases immediately 5’ and 3’ to each mutated base generating 96 possible mutation types (6 types of substitution ∗ 4 types of 5’ base ∗ 4 types of 3’ base). Mutational signatures are displayed and reported based on the observed trinucleotide frequency of the human genome. To extract de novo mutational signatures from mutation count matrix, non-negative matrix factorization was used. Known mutational signatures were searched from the COSMIC signature (v.2); totally 30 mutational signatures are available with additional information including the cancer types in which the signature has been found, proposed aetiology for the mutational processes underlying the signature, other mutational features that are associated with each signature and information that may be relevant for better understanding of a particular mutational signature [8].

### Functional analyses

To gain a functional annotation of genes harboring significant SNVs, Enrichr’s plugin [9] KEGG pathways [10] was used to identify relevant pathways.

### Statistical analysis

P value < 0.05 was considered as statistically significant. All statistical analyses were performed using the R software version 3.6.1 (R Foundation for Statistical Computing, Vienna, Austria).

## Results

As a result, a total of 143, 1,418 and 4,339 SNVs were found in Lymphoid, Myeloid and Fibroid group, respectively (Figure 2A and Supplementary Table 2-3). Large deviation in the count of SNVs among subgroups was thought to be due to difference of sample size. To evaluate impact of sample size bias on mutation characteristics, I checked mutation spectrum, which shows the relative contribution of each mutation type in the base substitution catalogs. As shown in Figure 2B, there was no clear difference of spectrum among samples, indicating difference of sample size might have little effect on further analysis.

**Figure 2.**
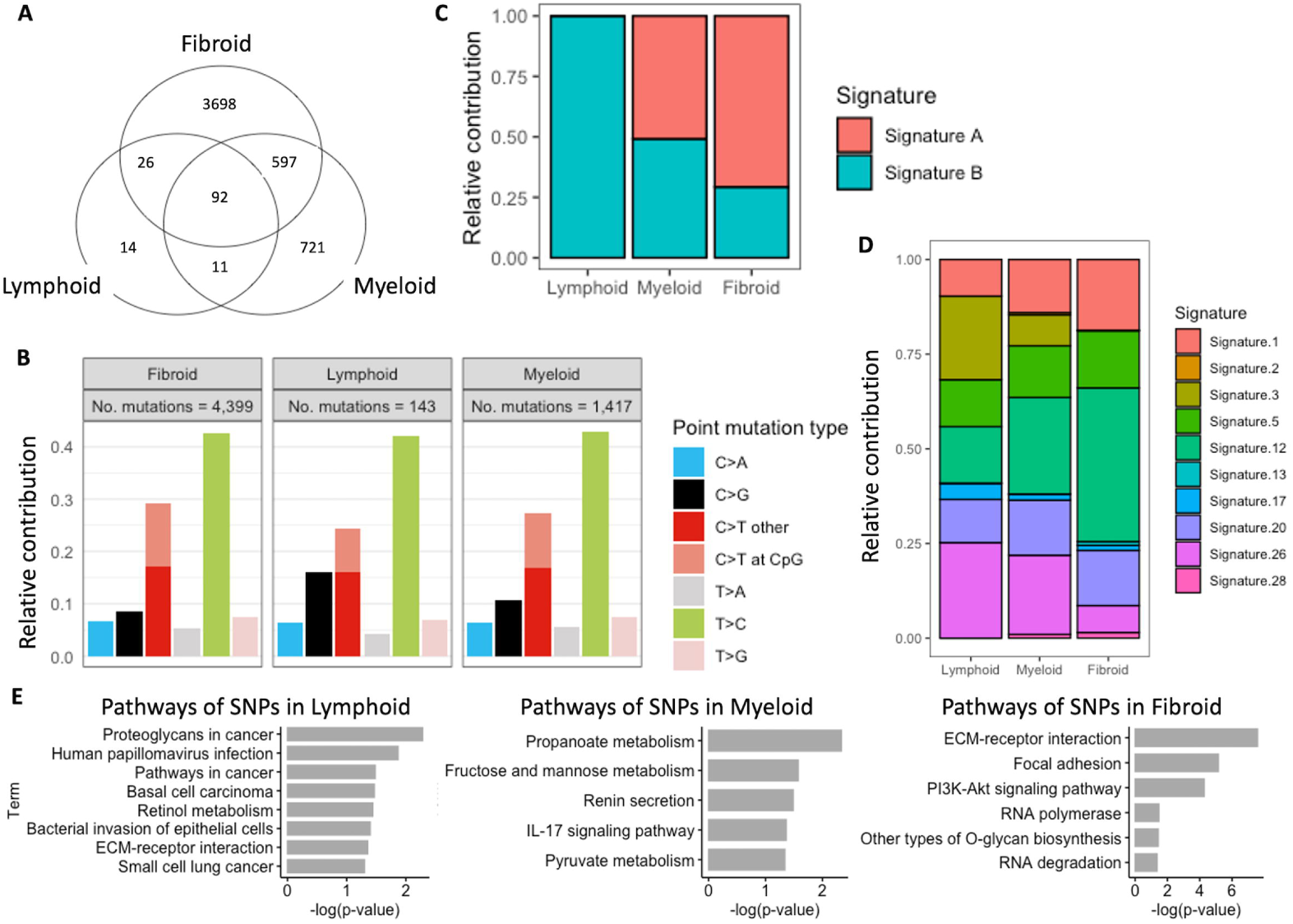
Landscape of Genetic mutations specific in synovitis. A) Venn diagram of SNVs in each pathotype group in patients with rheumatoid arthritis (Supplementary Table 2-3). B) Relative contribution of the nucleotide transition patterns according to each pathotype. C) De novo mutational signature characterized by a specific contribution of 96 base substitution types with a certain sequence context. D) Contribution of known mutational signatures (COSMIC signature, 1-30; https://cancer.sanger.ac.uk/cosmic/signatures_v2). To visualize the difference of contribution of each signature, signatures with small contribution were omitted. E) KEGG pathways based on specific SNVs in each pathotype (Supplementary Table 4-5). Only significant pathways (p value < 0.05) were shown. the catalogue f somatic mutations in cancer, COSMIC; single nucleotide variants, SNVs.

Then, I extracted mutational signatures which represent mutational processes (https://cancer.sanger.ac.uk/cosmic/signatures_v2). As with distribution of cell-lineage-specific transcripts [6], de novo mutational signature construction demonstrated mutational signatures were different according to histological subgroups, suggesting synovial tissue heterogeneity represents a divergent continuum of gene mutations with Fibroid at one end of the spectrum and Lymphoid at the other end of the spectrum (Figure 2C).

To investigate known characteristics of mutational signatures, I utilized COSMIC signature [8]. As well as de novo mutational signatures, signatures associated with *failure of DNA double-strand break-repair by homologous recombination* (Signature 3) and *defective DNA mismatch repair* (Signature 26) accumulated in order of synovium of Lymphoid, Myeloid and Fibroid group (Figure 2D). Conversely, Signature 1 and 12, which were enriched in synovium of Fibroid group, were reported as non-specific mutations in all cancer types and liver cancer-associated signature without known aetiology, respectively [8].

Finally, to annotate genes harboring significant SNVs which specifically existed in each histological group, pathway analysis was performed (Figure 2E and Supplementary Table 4-5). As a result, *Cancer-associated pathways, metabolic pathway*s and *cell adhesion pathways* were the most significantly enriched in Lymphoid, Myeloid and Fibroid group, respectively (Figure 2E). These results suggest variations of genetic mutations would contribute difference of inflammatory milieu in synovium.

## Discussion

In this study, accumulating gene mutations in inflamed synovium were identified in patients with early, pre-treatment RA. Consistent with expression of cell-specific gene modules [6], de novo mutational signatures reflected continuous relationship of synovial histology. Annotation analysis revealed defective DNA damage repair was associated with the level of inflammation in synovitis. Further, specific pathways in each histological group were thought to be aberrated according to pathway analysis based on genes harboring SNVs.

A large amount of RNA-seq data derived from lesion of autoimmune diseases have been generated in the last years. Subsequently, especially for gene expression profiling, several bioinformatics pipelines have been developed to investigate variants identification [5]. It allows for detection of previously unidentified variants that may be functionally important but difficult to capture using WGS or WES. Although our data didn’t include DNA sequencing data from the same samples that gave rise to the RNA-seq data, blood samples were used as non-lesional site control.

Signature 3 and 26 (associated with defective DNA damage repair) were enriched in order of Lymphoid, Myeloid and Fibroid. Considering level of inflammation according to pathotype, Signature 3 and 26 were thought to associate with high inflammation in synovium of RA. T cells, which were enriched in synovium of Lymphoid group [6], of human-synovium chimeric mice with inhibiting function of DNA damage repair showed tissue-invasive and pro-arthritogenic phenotype, and reconstitution of function alleviates synovitis, suggesting pathway of DNA damage repair could be therapeutic target for patients with lymphoid-rich synovitis [11].

Pathway analysis revealed accumulating SNVs in synovium represent different types of synovitis, with differing pathogenic process and inflammatory environment. Autoimmune diseases, including RA, have a clear association with cancer, though the strength of this association varies between different types of malignancies, and different populations [12]. *FZD6*, specific gene harboring SNVs in synovium of Lymphoid group (Supplementary Table 5), was involved in the development of chronic lymphocytic leukemia [13]. In addition, metabolic pathways, which was characteristic pathways in specific SNVs of Myeloid group, support macrophage functions as well as to sustain their polarization in specific contexts [14]. Moreover, cell adhesion pathways were enriched in genes of SNVs in Fibroid group. Integrins, which were found in SNVs of Fibroid group, were expressed in fibroblasts and endothelial cells in synovial tissue and remodel the extracellular matrix by inducing the expression of certain proteases [15]. Further study is needed to investigate impact of each mutation on these pathways.

This study suffers from several limitations. First, although there are a substantial number of genes with tissue-specific expression, part of synovium-specific variants, which were not expressed enough in blood, would also passed through the filter. To address this issue, additional DNA specimen of other than inflamed synovium tissue from same individual is needed. Second, other RNA-seq specific points could trigger false positive variants, such as RNA-editing sites and intron-exon junction sites [16,17]. Although the current study minimized these false positive errors by extracting only shared variants across individuals in each pathotype group, combination with other bioinformatics pipeline will be also useful [16,5].

In conclusion, mutational signatures represent different types of synovitis, supporting differing pathogenic process in early RA. To my knowledge, although single-cell RNA-seq of synovial biopsies would provide more complete genetic mutation at the level of individual cells and cannot exclude the possibility that they represent stage of the disease process, this is the first study demonstrating association between SNVs and types of synovitis. This data will promote our understandings about pathogenic status of synovitis, and suggests evidence that stratified treatments would be optimal according to histological subgroup. Further verification of gene mutations and functional analysis will require to validate significance of SNVs identified here.

## Data Availability

RNA-seq dataset used in the current study is deposited in ArrayExpress under Accession code E-MTAB-6141. Supplementary Table 2-5 are deposited at figshare (https://figshare.com/articles/Supplementary_Tables/11876553). All custom computer codes in the generation or processing of the described data are available upon reasonable request.

## Acknowledgement

Nothing to declare.

## Authors’ Contributions

All of conceptualization, formal analysis and writing were conducted by JI.

## Ethics approval

Not required because this study was conducted using only public database.

## Consent for publication

Not required.

